# Clinical and molecular characterization of the vinorelbine-platinum chemotherapeutic regimen in HER2(-) metastatic breast cancer

**DOI:** 10.1101/2024.06.19.24309211

**Authors:** I-Wei Ho, Yi-Ru Tseng, Chun-Yu Liu, Yi-Fang Tsai, Chi-Cheng Huang, Ling-Ming Tseng, Ta-Chung Chao, Jiun-I Lai

**Author notes:** Correspondence: Jiun-I Lai, M.D. Ph.D.

## Abstract

**Introduction:** Despite rapidly improving therapeutics, challenges remain in treatment of advanced breast cancer. Vinorelbine, a semisynthetic vinca alkaloid, is effective and well-tolerated in breast cancer treatment. The combination of vinorelbine and platinum-combination is a well-tolerated but underreported chemotherapy regimen. Bevacizumab, a VEGF-neutralizing antibody, has shown efficacy in HER2-negative metastatic breast cancer (mBC) when combined with chemotherapy. In this study we aim to investigate the clinical and molecular effects of vinorelbine-platinum in heavily pretreated HER2-negative mBC, as well as the role of addition of bevacizumab.

**Material and methods:** We conducted a retrospective study at Taipei Veterans General Hospital to evaluate the effectiveness of the vinorelbine-platinum regimen in heavily pretreated HER2-negative mBC patients from 2016 to 2020, with a portion of patients receiving additional bevacizumab. To model the molecular perturbations at a cellular level, transcriptional profiling of a triple negative breast cancer cell line treated with cisplatin-vinorelbine was done by RNA-sequencing.

**Results:** The cohort included 54 patients. 50% of the patients received ≥ 5 lines of systemic treatment in the metastatic setting. All the patients had received anthracyclines and taxane. In patients treated with vinorelbine-platinum combination, the median progression-free survival (PFS) and overall survival (OS) were 2.3 and 7.3 months, respectively. With bevacizumab, median PFS improved to 4.1 months. Objective response rate (ORR) and disease control rate (DCR) without bevacizumab were 11.1% and 27.7%, respectively, improving to 25% and 83.3% with bevacizumab. Adverse events occurred in 37.0% of patients, with no grade IV events reported. Transcriptional profiling revealed significant downregulation of MAPK pathway, angiogenesis, and growth factor signaling related genes.

**Conclusion:** The vinorelbine-platinum regimen, particularly with bevacizumab, shows efficacy even in heavily pretreated HER2-negative metastatic breast cancer patients. Molecular analyses of treated cells highlight potential targets and mechanisms of action, providing a basis for future therapeutic strategies.

## Introduction

Breast cancer is one of the most prevalent malignancies in the world, with more than 2.3 million newly diagnosed cases and 665,000 deaths globally in 2022.[1] Outcomes for breast cancer has improved drastically in the past 2 decades, with huge strides in all three major subtypes (Hormone receptor (HR) positive, HER2 positive, triple negative (TNBC)). Even in metastatic disease, overall survival reaches 4-5 years due to the advent of CDK4/6 inhibitors in HR(+) metastatic breast cancer (mBC)[2], dual blockade with pertuzumab plus trastuzumab[3]. With metastatic TNBC, overall survival (OS) reaches 2 years in PD-L1 high tumors with immunotherapy-chemotherapy combination [4], a huge improvement from prior-immunotherapy era, but nonetheless much worse outcome compared to HR(+) and HER2(+) subtypes. Overall, five-year survival rates in patient with metastatic breast cancers has been reported to be about 30% [5], although novel therapies are constantly improving this statistic. However, even with these highly efficacious regimens that prolonged overall survival, resistance eventually occurs, and in patients who have progressed on multiple systemic agents, therapeutic options are more limited.

Vinorelbine, a synthetic vinca alkaloid chemotherapeutic agent, disrupts microtubule formation during mitosis.[6] Due to its efficacy, well tolerability, and low toxicity or adverse effects, vinorelbine is commonly used in advanced metastatic breast cancer and is often the drug of choice in fragile or palliative settings.[7, 8] Vinorelbine is an integral component of metronomic therapy for breast cancer[9], and has even shown superior efficacy to weekly paclitaxel in a phase 2 trial [10].The role of platinum agents, such as cisplatin and carboplatin, is well established in breast cancer, particularly in TNBC. Platinum drugs are now recognized as standard of care in the neoadjuvant immunooncology treatment of non-metastatic TNBC, in the neoadjuvant setting without immunotherapy, and in metastatic TNBC (mTNBC) patients with BRCA1/2 mutations. [11–13] The combination of vinorelbine and platinum is a commonly used regimen in lung cancer[14], but its use in breast cancer is relatively less reported since 2 decades ago[15, 16]. This prompted us to ask whether vinorelbine plus platinum would be a tolerable regimen for heavily pretreated patients while maintaining acceptable toxicity profile.

Angiogenesis is recognized as a critical factor in the progression of metastatic breast cancer. [17] Elevated serum levels of vascular endothelial growth factor (VEGF) in patients with metastatic breast cancer have been associated with inferior clinical outcomes.[18] Bevacizumab, a VEGF neutralizing antibody, plays a critical role in inhibiting neoplastic angiogenesis, effectively impeding tumor progression.[19] Previous meta-analyses have established the efficacy of bevacizumab in treating HER2 negative metastatic breast cancer when used in conjunction with chemotherapy, resulting in improvements in progression-free survival (PFS) and objective response rate (ORR).[20] The addition of bevacizumab to chemotherapy in first line HER2 (-) metastatic breast cancer resulted in improved PFS (9.2 months) and response rate (49%) [21], and benefit was especially observed in poor prognosis patient groups including TNBC patients.

In our study, we aimed to investigate the clinical outcomes of platinum-vinorelbine regimen, especially in heavily pre-treated patients. In our cohort, 50% patients had received five or more lines of systemic therapy in the metastatic setting. We observed that the addition of bevacizumab increased PFS, response rate, and more importantly clinical benefit rate (CBR) to up to 83.3%, which is an important treatment goal in heavily pretreated patients. Transcriptional profiling revealed changes in gene expression profiles elicited by platinum-vinorelbine regimen, and interestingly revealed a potential synergy mechanism for bevacizumab. Our study provides molecular insight to complement clinical observations for a potentially useful regimen for a salvage chemotherapy setting.

## Material and methods

### Study Design and Setting

This retrospective cohort study was conducted at Taipei Veterans General Hospital in Taiwan, aiming to evaluate the efficacy of salvage chemotherapy regimens that involved the vinorelbine-platinum combination. The study also aimed to compare treatment responses, progression-free survival, overall survival, prognostic factors, and potential adverse outcomes. Specifically, the study focused on patients with HER2 negative metastatic breast cancer who underwent salvage chemotherapy between the years 2016 March and 2020 December. The investigation also explored the impact of incorporating bevacizumab into the treatment regimen. Data for this study were obtained from the electronic medical records. The extracted data encompassed a comprehensive range of information, including vital signs, symptoms, investigation procedures, workup details, diagnostic results, and patient management. Individuals lacking a confirmed diagnosis through histopathology verification or those referred to other hospitals for chemotherapy were excluded from the study. To define HER2 negativity, it was based on a HER2 result of 1+ or 0 in immunohistochemistry. In instances where the HER2 result was 2+, it was categorized as negative if paired with a negative outcome in the fluorescence in situ hybridization (FISH) test. Alongside chemotherapy details, we assessed age at breast cancer diagnosis, metastases sites, prior chemotherapy, treatment sequence, and adverse events. Institutional Review Board (IRB) approval for this research was secured from Taipei Veterans General Hospital (IRB TPEVGH No. 2023-09-007BC).

Overall response rate (ORR), disease control rate (DCR), progression-free survival (PFS) and overall survival (OS) were assessed according to the Response Evaluation Criteria in Solid Tumors (RECIST) v. 1.1 criteria. [22] Adverse events were also collected. Overall survival (OS) was defined as the period from the start of vinorelbine-platinum chemotherapy to the patient’s death from any cause. PFS was measured from the initiation of vinorelbine-platinum to the development of progressive disease (PD) or death.

### Statistical analysis

Statistical analysis was conducted using SPSS v26. All p-values were considered two-sided, and statistical significance was defined as p < 0.05. Survival curves were generated using the Kaplan-Meier method. Attributes related to the study were subject to univariate analysis and multivariate analysis to ascertain their association with PFS and OS. For both univariate and multivariate evaluations of prognostic factors pertaining to PFS and OS, the Cox proportional hazards model was applied. The outcomes were presented using hazard ratios (HR) as indicators of survival decline, along with their corresponding 95% confidence intervals.

### Cell culture and drug treatment

The hormone positive breast cancer cell lines MDA-MB-231 (purchased from ATCC Cell Lines (https://www.atcc.org/) were maintained in Dulbecco’s Modified Eagle Medium (DMEM, Gibco) respectively. All culture medium contained 10% Fetal Bovine Serum (FBS, Gibco) and 1% Penicillin Streptomycin (P/S, Gibco). Cells were incubated in 37°C, 5% CO2 under standard molecular biology conditions. Vinorelbine and cisplatin were purchased from Sigma-Aldrich (Missouri, United States) and diluted, stored per manufacturer instructions until use in cell culture.

### RNA-sequencing

MDA-231 cells (5 to 7 x 10^5^ per well) were seeded onto 6 cm dish and treated with 7μM cisplatin + 14μM vinorelbine or vehicle (DMSO) for 24 hours, washed in PBS, and RNA was extracted (Total RNA Isolation Kit, NovelGene). RNA was checked for quantity with a Nano-Drop (Thermo Fisher Scientific, RNA with A260/A280 ratios over 1.9 were used.) than stored at −80°C. Libraries were prepared using a TruSeq Stranded mRNA sample preparation kit (Illumina, San Diego, CA) from 500 ng of purified total RNA according to the manufacturer’s protocol in a reduced reaction volume. The finished cDNA libraries were assessed for quality using a Bioanalyzer and quantified with a Quant-iT dsDNA Assay kit (Thermo Fisher Scientific, Waltham, MA). The uniquely indexed libraries were multiplexed based on this quantitation and the pooled sample was quantified by qPCR using the Kapa Biosystems (Wilmington, MA) library quantification kit by the Molecular Biology Core Genomics Facility at the Dana-Farber Cancer Institute and sequenced on Illumina NovaSeq 6000 run with paired-end reads. Reads were processed to counts using the bcbio-Nextgen toolkit version 1.0.3a (https://github.com/chapmanb/bcbio-nextgen) as follows: (1) Reads were trimmed and clipped for quality control in cutadapt v1.12; (2) Read quality was checked for each sample using FastQC 0.11.5; (3) High-quality reads were then aligned into BAM files through STAR 2.5.3a using the human assembly GRCh37; (4) BAM files were imported into DEXSeq-COUNT 1.14.2 and raw counts TPM and RPKM were calculated. R package edgeR (McCarthy et al., 2012; Robinson et al., 2010) 3.18.1 (R version 3.2.1) was used for differential analysis and generate log fold change, p value and FDR.

The RNA-seq results were deposited into the Gene Expression Omnibus (GEO) database under the accession number GSE267621.

### Pathway analysis and data processing

Heatmaps, gene ontology (GO) analysis, were generated by the R packages “ClusterProfiler”, “enrichplot”, “ComplexHeatmap”. DAVID analysis was performed on the DAVID website (https://david.ncifcrf.gov/tools.jsp[23]).

## Results

### 1. Clinical characteristics of a HER2 negative metastatic breast cancer cohort

A total of 54 patients diagnosed with HER2 negative metastatic breast cancer and treated with the vinorelbine/platinum regimen were included in the study. The baseline characteristics of these patients are detailed in **Table 1**. The entire cohort comprised of female participants. The average age was 57.2 ± 8.7 years. Among them, 35 patients (64.8%) exhibited ER and/or PR positivity, while 19 patients (35.2%) were categorized as triple negative. More than 60% of patients had either bone, lung, or liver metastases, reflecting a cohort with heavy disease burden. All patients had received prior treatment with anthracyclines and taxane. 50% of all patients had previously received more than five lines of chemotherapy. 20 patients (37%) were de novo metastatic disease upon diagnosis, while 63% were recurrent disease. Other clinical details are described in **table 1**.

### 2. Clinical efficacy and adverse effects of the platinum plus vinorelbine, with or without addition of bevacizumab

**Table 2** presents the clinical efficacy data. The median progression-free survival (PFS) was recorded at 2.3 months, while the median overall survival (OS) was 7.3 months. With the addition of bevacizumab, median PFS was extended to 4.1 months (hazard ratio (HR) 0.54, p =0.04), and OS increased to 12.4 months. Among the patients, 11.1% (6/54) achieved a partial response (PR), 16.7% (9/54) exhibited stable disease (SD), and 72.2% (39/54) demonstrated progressive disease (PD). The calculated rates for objective response (ORR) and disease control (DCR) were 11.1% and 27.7%, respectively; and was increased to 25% and DCR to 83.3%, respectively. Interestingly, TNBC patients had a longer median OS (12 months), while HR(+) patients had an OS of 6.6 months.

Univariate analysis results are presented in **table 3**. Age > 65 and brain metastases were significantly associated with increased hazard ratio (HR) for PFS, while addition of bevacizumab significantly decreased HR for progression (HR=0.51, confidence interval (CI) 0.26-0.98, p=0.045). Age≥65, presence of lung metastasis, and prior treatment of ≥5 lines of chemotherapy were significantly associated with increased HR for overall survival. In our data, addition of bevacizumab did not significantly prolong OS.

The findings from the multivariate analyses, shown in **table 4**, indicated that the prior treatment of ≥5 lines of chemotherapy was significantly associated with both decreased OS and PFS. Additionally, lung metastasis was significantly associated with reduced OS.

Of the total 54 patients, 12 (22.2%) received additional bevacizumab treatment. Both univariate analysis (**Table 3**) and Kaplan-Meier plots (**Figures 1a** and **1b**) demonstrated a significant benefit in PFS associated with the addition of bevacizumab, although no significant improvement in OS was observed.

Within our cohort of 54 patients, ≥ grade 3 adverse events were documented in 20 individuals (37.0%), described in **Table 5**. Most of the adverse effects were related to bone marrow toxicity, including grade III neutropenia accounted for 25.9% of cases, and grade III anemia in 12.5%. Importantly, no grade IV adverse events were reported in the study.

### 3. Transcriptional profiling of triple negative breast cancer cells treated with cisplatin-vinorelbine reveals molecular insights of cytotoxic mechanisms

To model the molecular perturbations by cisplatin-vinorelbine combination, we treated the triple negative breast cancer MDA-MB-231 cell line with a combination of cisplatin plus vinorelbine, in comparison to vehicle treatment. Total RNA extracted from treated cells underwent RNA-sequencing for transcriptional profiling. Differential expression genes (DEGs) were analyzed for enriched pathways. We observed that in gene ontology (GO) analysis, cisplatin-vinorelbine combination resulted in strong impact of growth factor binding, PDGFR binding, TGF-beta binding, as well as other pathways (**Fig 2A**). In pathway analysis by DAVID, the top enriched pathways were the MAPK pathway and the Ras signaling pathway (**Fig 2B**). Heat map analysis of the differentially expressed genes from the MAPK pathway revealed downregulation in several growth factors, including EGF, CSF1, NGFR1, TGFA, VEGFA, and CSF1R (**Fig 2C**), suggesting that platinum-vinorelbine regimen resulted in transcriptional suppression of many tumors related growth factor signaling pathways.

## Discussion

Despite rapidly increasing therapy options in metastatic breast cancer, treatment options for heavily pretreated patients are limited and clinical efficacy is usually modest. In a real-world study using the Flatiron electronic database, the researchers used the criteria for MONARCH 1 to investigate outcome in a highly pretreated setting[24]. The investigators selected female HR (+) HER2(-) mBC patients who received monotherapy using eribulin, vinorelbine, gemcitabine, capecitabine. These patients had an ECOG performance status of 0-1. In this cohort containing 108 patients, median OS was 13.6 months. In a retrospective study of 78 prior treated mBC (> 2 lines) patients who received eribulin monotherapy, the median PFS was 3 months, and median OS was 7 months [25]. This study has efficacy data more similar to our study, in a cohort of heavily pretreated patients with modest survival outcomes but acceptable toxicity profile and tolerability. In patients heavily pretreated or unfit for intensive chemotherapy, metronomic chemotherapy is often employed and offers modest response rates and survival outcomes, while offering improved clinical benefit rate (CBR)[26].

To date, the optimal first-line regimen for metastatic TNBC consists of chemotherapy with or without immunotherapy or PARP inhibitors, depending on PD-L1 expression and germline BRCA mutation status. Following the failure of first-line therapy, sacituzumab govitecan is has shown benefit as later line treatment.[27, 28] However, besides Sacituzumab govitecan, there is no clear consensus on the optimal chemotherapy regimen for metastatic TNBC. Eribulin, capecitabine, ixabepilone, and platinum-based therapies are all considered reasonable options, yet no single regimen is regarded as superior in this setting.[27] Currently, metastatic TNBC remains incurable. This area of research represents a significant unmet clinical need, with novel therapeutic agents and combinations urgently required. In our study, we observed that although PFS was similar between TNBC and HR(+), OS was markedly increased in TNBC (12 months). Although the sample size is small, our study provides interesting insights that is in line with previous reports that addition of bevacizumab may be beneficial in metastatic TNBC. In the ATHENA study, the TNBC subgroup when receiving 1^st^ line bevacizumab plus chemotherapy had a time to progression of 7.2 months and OS of 18.3 months, with an overall response rate of 49% [29]. Complemented with our RNA-seq data, our study suggests that bevacizumab addition should be considered in TNBC patients for achieving better disease control status.

The combination of vinorelbine and platinum has demonstrated efficacy against TNBC in previous observational studies, even in heavily pretreated patients. [30, 31] Our study extends these findings, showing efficacy in both TNBC and hormone receptor-positive patients, with an objective response rate (ORR) of 11.1% and a disease control rate (DCR) of 27.7%. Furthermore, co-administration with bevacizumab enhanced these outcomes, elevating the ORR to 25% and the DCR to 83.3%. interestingly, both univariable and multivariable analyses consistently pointed towards patients who had undergone 5 or more lines of chemotherapy experiencing reduced PFS and OS. This aligns with earlier research, which demonstrated a decline in clinical benefit rate from 85% in the initial line to 54% by the fourth line of treatment.[32] It is noteworthy that the response rate in our study was lower compared to prior studies. [30, 31], but it must be taken into account that half of the patients in our cohort received more than five lines of prior systemic therapy. Considering the heavily treated nature of our cohort, it remains impressive that in such frail and refractory patient group, the addition of bevacizumab to vinorelbine/platinum could improve the response rate to 25%, with a very high disease control rate. Our cohort, although with small sample size, suggests a promising combination especially in late stage, refractory patients.

The MAPK signaling pathway plays a crucial role in breast cancer development, particularly influencing the expression of estrogen receptor (ER), progesterone receptor (PR), and HER2. It is closely associated with the invasion, metastasis, and prognosis of TNBC.[33] Elevated MAPK activity correlates with shorter survival times in TNBC patients, suggesting its potential as a prognostic indicator.[33, 34] Various mitogens such as TGF-α, EGF, VEGF, and PDGF-β bind to their respective receptors, triggering RAS activation, which subsequently stimulates the MAPK pathway.[35] In our RNA sequencing analysis of triple-negative breast cancer cell lines treated with a cisplatin-vinorelbine combination, we observed downregulation of the MAPK pathway. Interestingly, VEGF-A was shown to be upregulated after cisplatin/vinorelbine treatment. This suggested a rationale for combination therapy with cisplatin/vinorelbine and bevacizumab, enhancing the VEGF-A targeting efficacy of bevacizumab. More preclinical and clinical studies will shed light on this issue, whether VEGF-A serum concentrations are correlated with treatment outcome of this regimen, and whether anti-angiogenesis activity is impaired. Notably, targeting various steps in the MAPK pathway with BRAF plus MEK inhibitors has been established as the standard of care for patients with advanced-stage melanoma harboring BRAF V600 mutations.[36] MEK inhibitor Trametinib has been proposed to possess activity towards TNBC in phase 2 study.[37] We hypothesize that combining this approach with bevacizumab could yield synergistic antitumor effects through vertical inhibition. Interestingly, our RNA-seq data revealed that many of the aforementioned growth factors pathways are downregulated by treatment of platinum-vinorelbine, suggesting promising clinical activity for further consideration.

It is important to acknowledge that our study had a limited sample size, thus drawing definitive conclusions regarding the specific characteristics for the vinorelbine-platinum regimen is challenging. Further investigations with larger cohorts are warranted to pinpoint which patient groups can derive significant benefits from the vinorelbine-platinum regimen. However, we believe that our study provides valuable insights into a cohort with an extremely poor prognosis and limited clinical options, contributing preliminary data that could guide further development of systemic therapies. Given that vinorelbine targets microtubules, other anti-microtubule agents such as eribulin, ixabepilone, and taxane also play significant roles, not only in first-line but even in heavily pretreated patients.[27, 38, 39] It is intriguing to speculate that newer agents, such as antibody-drug conjugates with tubule-targeting payloads like enfortumab vedotin, may demonstrate clinical activity in metastatic TNBC, with or without the addition of bevacizumab.[40]

## Conclusions

The vinorelbine/platinum regimen, particularly when combined with bevacizumab, demonstrates efficacy in heavily pretreated HER2-negative metastatic breast cancer patients. Adverse events associated with this treatment were manageable. Molecular analyses of treated cells have identified potential targets and mechanisms of action, laying the groundwork for the development of future therapeutic strategies.

## Figure legends

Table 1: Clinical characteristics of patients receiving platinum-vinorelbine regimen

Table 2: Clinical outcomes of patients receiving platinum-vinorelbine regimen

Table 3-4: univariate (Table 3) and multivariate (Table 4) analysis of relevant clinical factors for PFS and OS

Table 5: prevalence of adverse effects with platinum-vinorelbine regimen

Fig 1A-B: Kaplan Meier plots of all patients are shown for a) progression-free survival with/without Bevacizumab b) overall survival with/without Bevacizumab

Fig 2A: gene ontology (GO) analysis of platinum-vinorelbine regimen treatment in TNBC cells

Fig 2B: DAVID analysis of RNA-seq differentially expressed genes (DEG)

Fig 2C: heatmap comparing differentially changed genes in the MAPK pathway between platinum-vinorelbine treated cells and DMSO treated cells. vin_cis: vinorelbine plus cisplatin treated cells. 2 biological repeats were done for each treatment condition.

## Data Availability

All data produced in the present study are available upon reasonable request to the authors
All data produced in the present work are contained in the manuscript

---

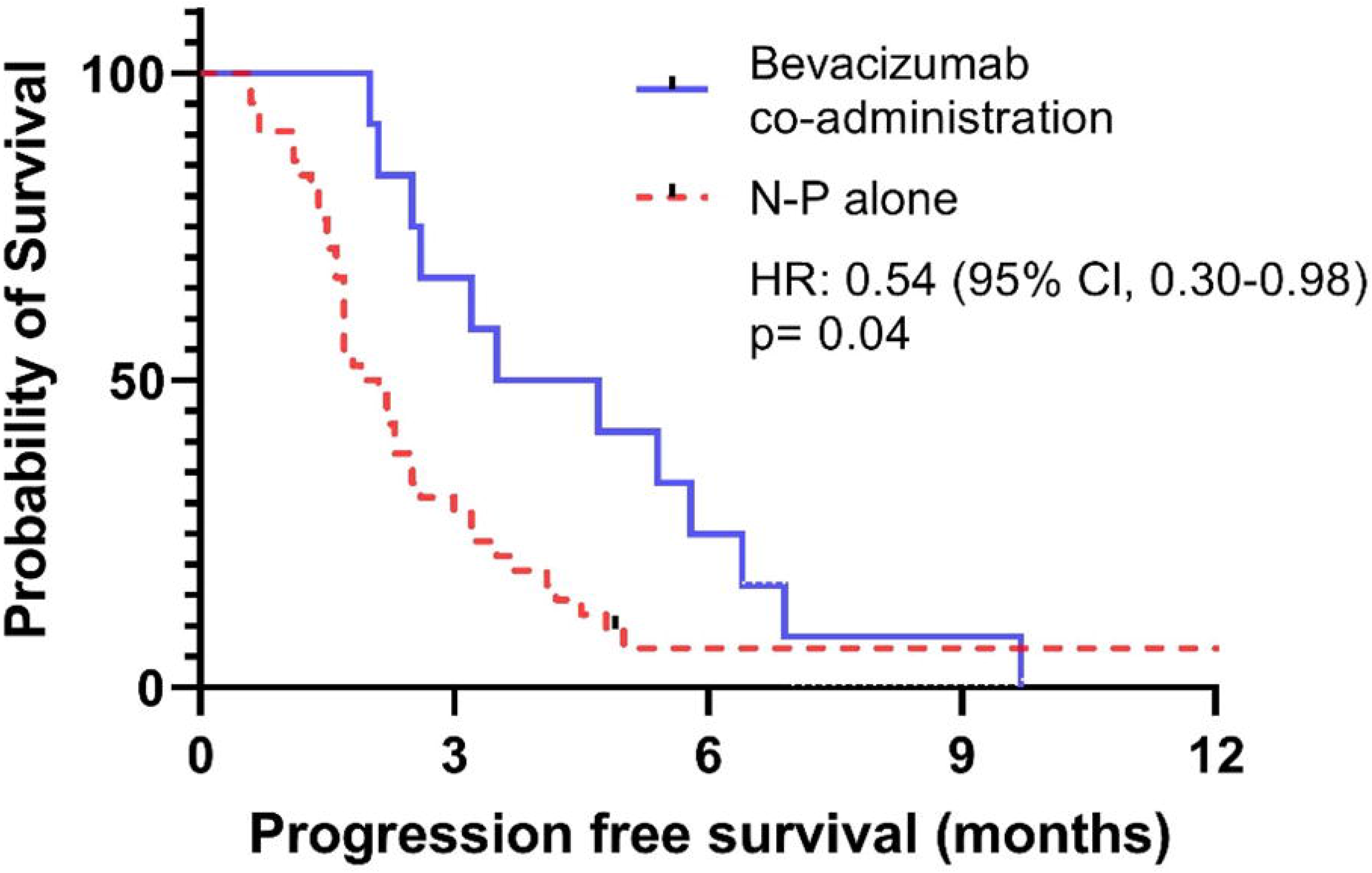

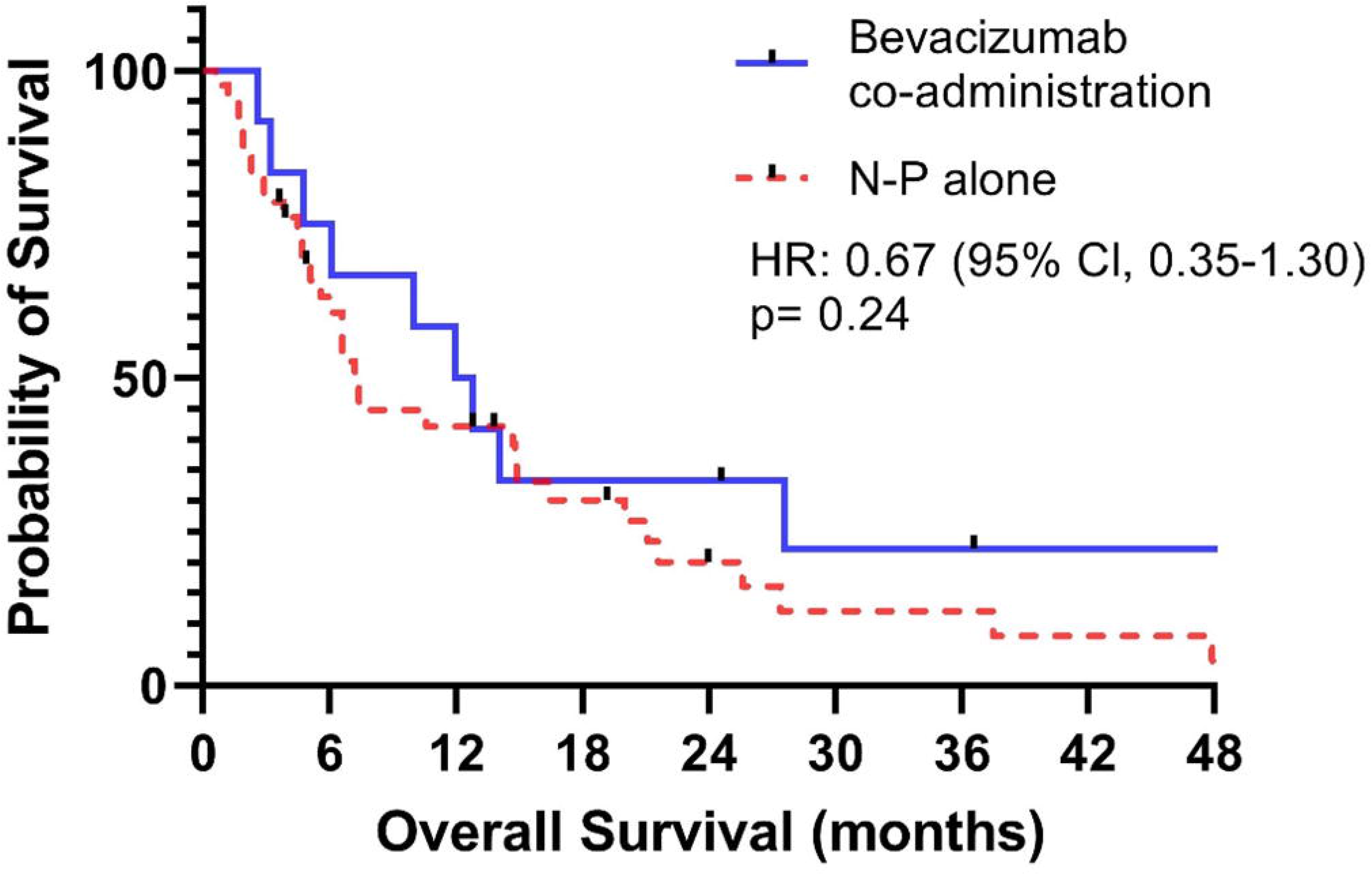

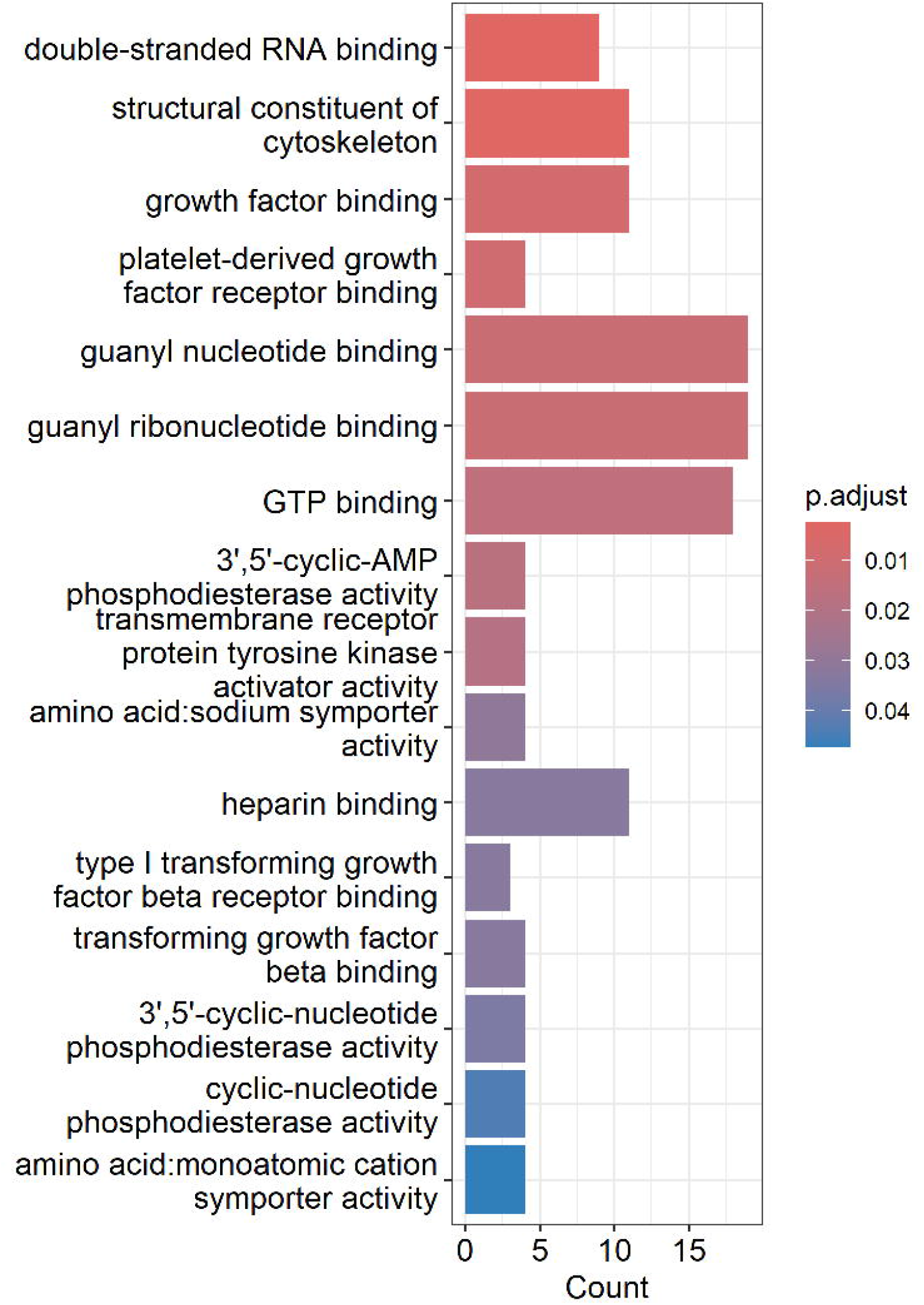

| Term | Count | P Value | Fold Enrichment | FDR |
| --- | --- | --- | --- | --- |
| MAPK signaling pathway | 29 | 7.39E-06 | 2.568 | 6.8E-04 |
| Ras signaling pathway | 24 | 2.36E-05 | 2.710 | 1.3E-03 |
| Rap1 signaling pathway | 21 | 1.11E-04 | 2.665 | 4.7E-03 |
| Lipid and atherosclerosis | 18 | 0.003 | 2.231 | 4.1E-02 |
| Legionellosis | 8 | 0.005 | 3.807 | 5.7E-02 |
| Alcoholic liver disease | 13 | 0.007 | 2.440 | 7.0E-02 |

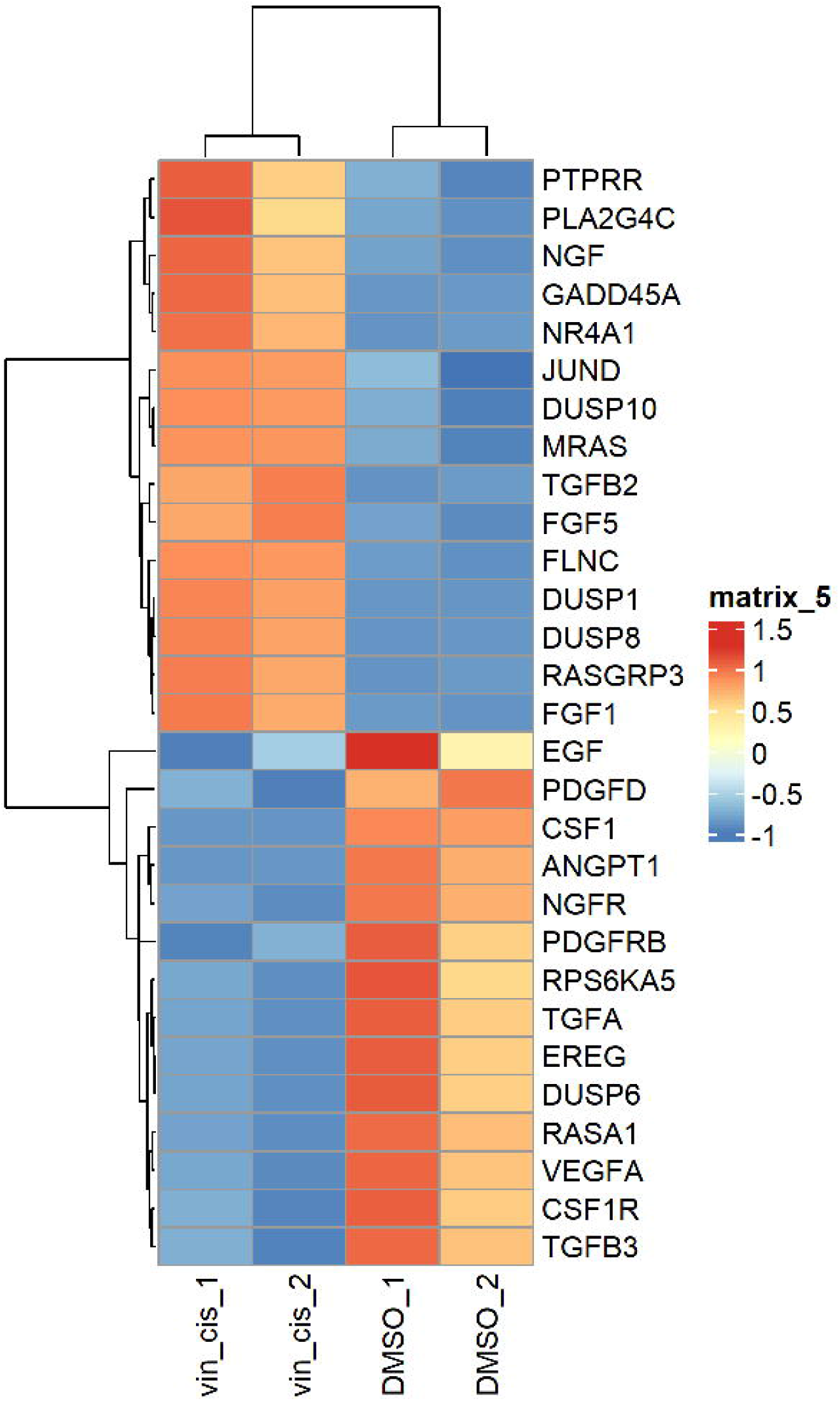

